# A Novel Maxillofacial Technology for Drug Administration-A Randomized Controlled Trial Using Metronidazole

**DOI:** 10.1101/2024.09.11.24313269

**Authors:** Sajani Ramachandran, Ravichandran Kandasamy, Kavita Verma, Jayahshri Murugan, Jishnu Sudhakar, Hridwik Adiyeri Janardhanan, U.R Anoop

## Abstract

**Aim:** To compare the efficacy of a novel maxillofacial drug delivery technology with that of oral administration by analyzing the presence of administered metronidazole in plasma.

**Material & Methods:** The patients reporting to the Dental outpatient department with acute pulpitis in relation to maxillary molar were examined and recruited for the study. All consenting patients fulfilling the inclusion criteria during the study period were included.

The selected patients were randomly divided into two groups using computer generated sequence of random numbers.

**Group I**: In the control group patients, a single dose of 400 mg of metronidazole in the form of a tablet was administered through the oral route, after biomechanical preparation of root canals.

**Group II**: In the experimental group patients, 5mg of metronidazole in the form of an infusion solution was administered into the pulp cavity of a maxillary molar using the maxillofacial route and technology, after biomechanical preparation of root canals.

The blood samples were collected at 15 minutes and 30 minutes after administration of metronidazole. The plasma samples were then analysed for the presence of metronidazole using high performance liquid chromatography (HPLC).

**Results:** Out of ten, 5 (50%) samples in the oral route group and all 9 (100%) samples in the maxillofacial route group had the presence of the drug (p = 0.033). There was no statistically significant difference in presence of the drug between 15 minutes and 30 minutes in the oral route (p = 1.0) and in the maxillofacial route (p = 0.687). Further, the mean value of the area under the HPLC curve after 15 minutes was found to be similar in both the groups (p = 0.4). At 30 minutes also the area under the HPLC curve between the groups was not statistically significant (p = 0.156)

**Conclusion:** The maxillofacial drug delivery technique can be an effective alternate route for painless and controlled drug delivery.

## Introduction

Drug delivery techniques are pivotal for providing effective treatments for patients by maximizing targeted drug delivery and minimizing side effects ^1, 2^. Drug delivery is challenging because of biological and physiological barriers. Existing routes of drug administration include enteral, parenteral, transdermal, nasal, pulmonary, ocular and vaginal routes ^3^. As the clinical use of nucleic acids, proteins and antibodies as drugs is increasing, the need for newer modes of drug delivery becomes crucial ^4, 5^. A drug delivery technology which can provide painless, repeated and controlled drug administration for different types of drugs holds immense potential.

In this study, we present a novel maxillofacial drug delivery technology and compare the efficacy of maxillofacial administration of metronidazole with that of oral administration of metronidazole.

## Objectives

1. To determine the quantity of metronidazole in the plasma at 15 minutes and 30 minutes interval when delivered through maxillofacial route
2. To determine the quantity of metronidazole in the plasma at 15 minutes and 30 minutes interval when given orally
3. To compare the plasma level of orally administered metronidazole with that of maxillofacial administration at 15 minutes and 30 minutes.

## Material & Methods

The study was conducted after getting clearance from the Institutional Ethics Committee and after registration of the study prospectively with the clinical trials registry – India. Patients reporting to the Dental outpatient department with acute pulpitis in relation to maxillary molar were examined and recruited for the study. All the consenting patients fulfilling the inclusion criteria during the study period were included.

### Inclusion Criteria

1. Patients reporting with pulpitis.
2. Consenting adult patients above the age of 18 years.
3. Patients requiring root canal treatment for maxillary 1st molar.
4. Patients with dental caries with pulp exposure.
5. Patients with no contra-indications for root canal procedure.
6. Patients who have no known systemic problems and are not on regular medications.

### Exclusion Criteria

1. Patients with periapical lesions not involving the pulp.
2. Patients with known allergy to metronidazole.
3. Patients who have taken metronidazole or any other drugs for the past 72 hours.
4. Patients who have bleeding disorders, liver and kidney problems or are on anti-platelet drugs.

### Sample Size Determination

To achieve an effect size of 1.3 between the two groups for the plasma value of metronidazole (i.e., in those for whom drug was administered orally and in those for whom drug was administered using the maxillofacial drug delivery technology) with 5% level of significance and 80% power, the sample size required per group was estimated as nine. Accounting for 10% attrition it was increased to 10 per group. Effect size was estimated at 1.3 from Stolz K et al ^6^.

### CONSORT diagram

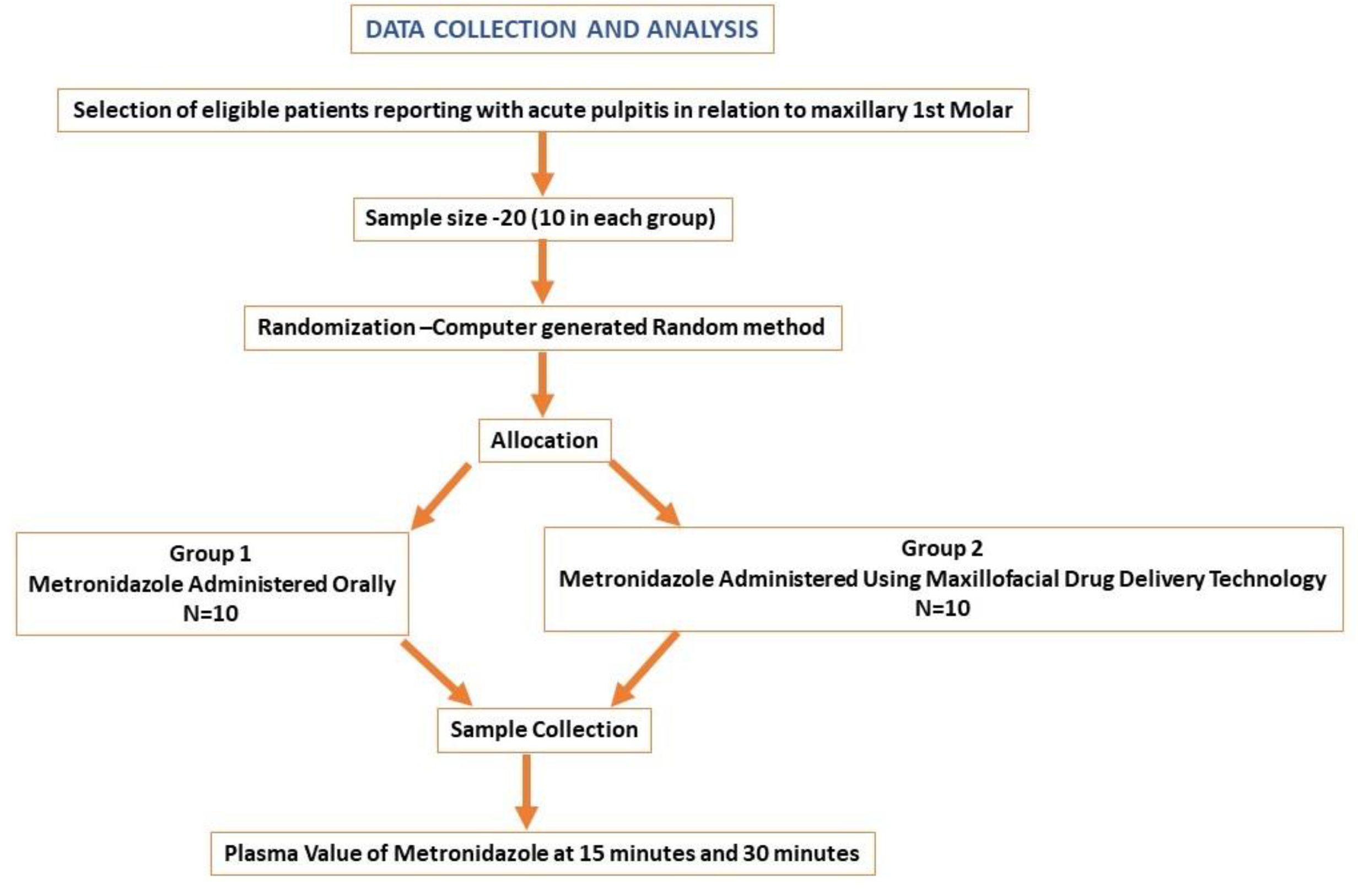

The selected patients were randomly divided into two groups of 10 patients each using computer generated sequence of random numbers.

Group I: The control group comprised of patients in whom 400 mg of metronidazole was given orally after biomechanical preparation of root canals.

Group II: The experimental group comprised of patients in whom 5mg of metronidazole was administered into the pulp cavity of a tooth using the maxillofacial technique after biomechanical preparation of root canals.

Carious upper molars with involvement of pulp were selected and root canal treatment was initiated under local anaesthesia. After opening the access to the pulp chamber, the pulp canals were located and the pulp was extirpated. Biomechanical preparation of the root canals was done with rotary files and irrigation was done with saline.

In Group 1, the access opening to the pulp cavity was closed with zinc oxide eugenol paste and each patient was given one dose of 400 mg of metronidazole orally in tablet form.

In group II after the extirpation of pulp and widening of the pulp canals, the canals were irrigated with saline and dried. Tooth preparation was done and a preliminary impression was made for a temporary crown. The crown was then fabricated for use with the maxillofacial drug delivery technology. The crown coupled to the maxillofacial drug delivery system was temporarily secured to the tooth and metronidazole infusion solution containing 500 µg/100 µl was delivered into the pulp cavity in a controlled manner for 10 minutes at the rate of 100µl/min. After drug administration, the tooth and access to the pulp cavity was closed with zinc oxide eugenol cement.

#### Blood sample collection

The blood samples of both the groups were collected prior to any intervention. Following the drug administration the blood sample (3ml) of both the groups were collected at 15 minutes and 30 minutes by an experienced technician using sterile vacutainer EDTA tubes. The plasma was isolated by centrifuging at 3000 rpm for 5 minutes in the institution lab. The isolated plasma was stored at-80°C till dispatch. The collected plasma was dispatched by using appropriate dry ice compatible shipping boxes to the HPLC lab for analysis.

### Statistical Analysis

Mean (± standard deviation) and median with inter quartile range were used to describe the continuous variables, while frequencies and percentages were used for categorically measured variables. The Kolmogorov–Smirnov test was applied to test the assumption of normality. Fisher’s exact / McNemar test was used to assess the statistical significance of binary outcome, appropriately. Wilcoxon Signed Ranks test / Mann-Whitney Test was applied to the continuously measured dependent outcome variable. The SPSS IBM statistical computing software, version 29.0, was used for the statistical data analysis. The statistical alpha significance level was considered to be at the 0.050 level.

## Results

Though 10 patients were recruited in both the groups, the samples of one patient in Group II could not be analysed by HPLC as those samples were turbid. Hence this sample was excluded and samples of only 9 patients in this Group were analysed by HPLC.

The presence of Metronidazole in the plasma was analyzed in both the groups. In the group where the medication was given orally, 5 (50%) samples had metronidazole where as in the maxillofacial route, 9 (100%) of the samples had the presence of the drug. This was found to be statistically significant (p value 0.033) [Table1].

**Table 1.** Comparing the presence of metronidazole between the oral route and maxillofacial route.

| Status | Oral tablet group | Maxillofacial route group | p value |
| --- | --- | --- | --- |
| Present | 5 (50.0) | 9 (100.0) | 0.033 |
| Absent | 5 (50.0) | 0 (0.0) |  |

**Note: numbers in bracket are percentage**

On comparing the presence of metronidazole administered through the oral route at 15 minutes and 30 minutes, the presence of the drug was noticed in 4 (40%) samples, whereas at 30 minutes, with addition of one more sample, the presence of drug was found in 5(50%). This was not found to be statistically significant p value 1.0 [Table2].

**Table 2.** Comparing the presence of Metronidazole at different point in time within the oral tablet group.

| Time/ Status | 30 Minutes |  | Total | p value |
| --- | --- | --- | --- | --- |
|  | Present | Absent |  |  |
| 15 Minutes |  |  |  |  |
| Present | 4 | 0 | 4 | 1.000 |
| Absent | 1 | 5 | 6 |  |

In the maxillofacial route, at 15 minutes presence of drug was observed in 5 (55.6%) samples and another 4 (44.4%) samples showed the presence of the drug at 30 minutes. However, out of the 5 seen at 15 minutes, in 2 samples presence of the drug was not observed at 30 minutes. This was found to be statistically insignificant (p value 0.687) [Table 3]

**Table 3.** Comparing the presence of Metronidazole at different point in time within the maxillofacial route group.

| Time/ Status | 30 Minutes |  | Total | p value |
| --- | --- | --- | --- | --- |
|  | Present | Absent |  |  |
| 15 Minutes |  |  |  |  |
| Present | 3 | 2 | 5 | 0.687 |
| Absent | 4 | 0 | 4 |  |

The mean value of area under the HPLC curve for metronidazole administered through oral route at 15 minutes was 12, 460.9 (± 28,692.8) AU*min and at 30 minutes was 39,227.0 (± 93,563.8) AU*min. This was not statistically significant (p value 0.08). The mean value of area under the HPLC curve for metronidazole administered through maxillofacial route at 15 minutes was 1, 31,902.1 (± 3, 58,584.3) AU*min and at 30 minutes was 2, 34,461.6 (± 5, 69,942.9) AU*min. This also was not statistically significant (p value 0.374) [Table 4].

**Table 4.** Distribution of area under the HPLC curve at different time points in both the group.

| Parameters/ Time | Oral tablet group | Maxillofacial route group |
| --- | --- | --- |
|  | AU*min | AU*min |
| 15 Minutes |  |  |
| Mean (± SD) | 12,460.9 (± 28,692.8) | 1,31,902.1 (± 3,58,584.3) |
| Median (IQR) | 0 (10,358.8 – 0) | 11,371.0 (37,328.5 – 0) |
| 30 Minutes |  |  |
| Mean (± SD) | 39,227.0 (± 93,563.8) | 2,34,461.6 (± 5,69,942.9) |
| Median (IQR) | 1,201 (40,947.8 – 0) | 38,863 (1,21,008.5 -7,469.0) |
| p value | 0.080 | 0.374 |

**IQR: Inter Quartile Range**

On further analysis, the mean value of area under the HPLC curve after 15 minutes was found to be similar in both the groups and statistically not significant (p value 0.4). At 30 minutes also area under the HPLC curve between the groups was not statistically significant (p value 0.156) [Table 4]

## Discussion

Novel drug delivery technologies have become game changers in clinical practice with the advent of newer drug formulations in the form of nucleic acids, peptides, proteins and antibodies. Novel drug delivery technologies and newer routes for drug administration are now crucial for delivering drugs across challenging biologic and physiologic barriers.

The existing routes of drug delivery like the enteral, parenteral, transdermal, nasal, pulmonary, ocular and vaginal routes have limitations ^3^. Therefore, there is a need for a drug delivery technology and a route which can provide painless, repeated and controlled drug administration for different types of drugs at low doses.

In this study, we present a novel maxillofacial technology and a new route for administering low doses of a drug. Further we compare the efficacy of the maxillofacial route of administration with that of oral route of administration. An infusion solution of metronidazole containing 500µg/100µl metronidazole was administered into the pulp cavity of a maxillary molar using a novel maxillofacial drug delivery technology at the rate of 100µl/min and the presence of the drug was assessed in the plasma using HPLC. It was compared with oral administration of 400mg of metronidazole in tablet formulation. The low dose delivered using the maxillofacial technology is an advantage when compared with the high dose that is used for oral administration.

Studies have shown that the minimum inhibitory concentration of metronidazole for most anaerobes including Bacteroides fragilis is less than 6µg/ml ^7, 8^. After an oral dose of 250mg, the peak serum level of the drug was found to be 6.2µg/ml and after an oral dose of 500 mg, the peak serum level was 11.2µg/ml. Similarly, after a single oral dose of 2gm, the peak serum level was 40µg/ml ^9^.

In this study, metronidazole was chosen for administration because the study samples comprised patients requiring root canal treatment. Infection of the root canal system is polymicrobial. Intracanal medicaments are used in infected root canals to eliminate remaining bacteria after root canal instrumentation ^10^.

Metronidazole, a nitroimidazole group of drugs with broad antibiotic spectrum is effective against obligate anaerobic bacteria. It is bactericidal for anaerobic organism at very low concentrations of 0.5%. The advantage of using metronidazole in dentistry as an intra canal agent includes ready availability, rapid bactericidal action, good penetration, cost effectiveness, acceptable pharmacokinetics and pharmacodynamics, undiminished antimicrobial activity and inability of susceptible organisms to develop resistance. There is no problem with readministration of the metronidazole ^10^. On evaluating the antibacterial activity of 0.12% chlorhexidine gel, 10% metronidazole gel, calcium hydroxide plus distilled water, calcium hydroxide along with camphorated para-monochlorophenol and calcium hydroxide along with glycerine using an agar diffusion test, it was found that metronidazole caused inhibition of the growth of all the tested obligate anaerobes ^11^. Further when metronidazole was used as an intra canal medicament in infected primary teeth, the success rate was 83% ^12^. Metronidazole when used as intra-canal medicament in infected primary molars had a success rate of 85% after 24 months ^13^.A study for evaluating the efficacy of metronidazole gel versus metronidazole solution against Enterococcus faecalis in abscessed primary molar found Metronidazole gel (3 % w/v) to be more effective than metronidazole solution (0.5 % w/v) against E. faecalis ^14^.

10 participants in oral route group and 9 patients in maxillofacial route group were considered for analysis. The presence of metronidazole in the plasma was analyzed in both the groups. In the group where the medication was given orally, 5 (50%) samples had metronidazole where as in the maxillofacial route, 9 (100%) of the samples had the presence of the drug. This was found to be statistically significant (p value 0.033) [Table1]

The maxillofacial drug delivery technique can be an effective alternate route for painless and controlled drug delivery.

## Data Availability

All data produced in the present study are available upon reasonable request to the authors

## Statements and Declarations

### Ethics Approval

The Institutional Ethics Committee of Pondicherry Institute of Medical Science gave ethical approval (IEC: RC/2022/169) The study has been registered prospectively with ICMR Clinical Trial Registry - India as CTRI/2023/03/050279.

## Data Availability

Clinical data is available upon reasonable request to the corresponding author.

## Acknowledgment

The authors acknowledge Dr. D. Saravanan, Ph.D., Coordinator, and Ms. S. Kavitha, M.Pharm., HPLC Analyst, from the National College Instrumentation Facility, Tiruchirapalli, India, for their invaluable support in completing the HPLC study.

